## Supplementary Material for "Attenuated viral strains of priority pathogens for potential use in controlled human infection model studies: A scoping review"

D.O. Hamilton^1^, V. Simpson^1^, T. Fox^1^, V. Lutje^1^, A. Kohl^2^, D. M. Ferreira^1,3^, B. Morton^1^

1. Department of Clinical Sciences, Liverpool School Tropical Medicine, Liverpool, UK

2. Centre for Neglected Tropical Diseases, Departments of Tropical Disease Biology and Vector Biology, Liverpool School of Tropical Medicine, Liverpool, UK

3. Oxford Vaccine Group, Department of Paediatrics, University of Oxford, Oxford, UK.

### Supplementary material

## S1

PRISMA-Scr Checklist – see separate document

### S2 Search strategy

Ovid MEDLINE(R) ALL <1946 to March 27, 2024>

1	Ebola virus.mp. or Ebolavirus/

2	Hemorrhagic Fever, Ebola/ or ebola.mp.

3	1 or 2

4	Lassa virus.mp. or Lassa virus/

5	Lassa fever.mp. or Lassa Fever/

6	4 or 5

7	Nipah virus.mp. or Nipah Virus/

8	Henipavirus Infections/

9	7 or 8

10	Rift Valley Fever/

11	Rift Valley Fever virus.mp. or Rift Valley fever virus/

12	10 or 11

13	Middle East Respiratory Syndrome Coronavirus/ or Middle eastern respiratory.mp.

14	(MERS* or MERS-CoV).mp.

15	13 or 14

16	Chikungunya Fever/ or Chikungunya virus/ or Chikunguya.mp.

17	pseudochallenge*.mp.

18	((attenuated or mutant or mutated) adj2 (virus* or pathogen* or strain*)).mp.

19	(whole genome adj2 (vaccin* or immun* or challenge*)).mp.

20	(experimental adj3 (vaccin* or immuni* or challenge*)).mp.

21	((preclinical or pre-clinical) adj2 study).mp.

22	clinical trials as topic/ or clinical trials, phase i as topic/ or clinical trials, phase ii as topic/ or clinical trials, phase iii as topic/ or clinical trials, phase iv as topic/

23	placebo*.mp.

24	controlled trial.mp.

25	CHIM.mp.

26	controlled human infection.mp.

27	human challenge.mp.

28	Vaccines/

29	Vaccination/

30	(((nonhuman or non-human) adj2 primate*) or NHPs).mp.

31	17 or 18 or 19 or 20 or 21 or 22 or 23 or 24 or 25 or 26 or 27 or 28 or 29 or 30

32	3 and 31

33	6 and 31

34	9 and 31

35	12 and 31

36	15 and 31

37	16 and 31

Embase 1947-Present, updated daily

1	Ebola virus.mp. or Ebolavirus/

2	Ebola hemorrhagic fever/ or ebola.mp.

3	1 or 2

4	Lassa virus.mp. or Lassa virus/

5	Lassa fever.mp. or Lassa Fever/

6	4 or 5

7	Nipah virus.mp. or Nipah Virus/

8	Rift Valley Fever/

9	Rift Valley Fever virus.mp. or Rift Valley fever bunyavirus/

10	8 or 9

11	Middle East Respiratory Syndrome/ or Middle eastern respiratory.mp.

12	MERS.mp.

13	11 or 12

14	Chikungunya virus/ or Chikunguya.mp.

15	pseudochallenge*.mp. or virus attenuation/

16	((attenuated or mutant or mutated) adj2 (virus* or pathogen* or strain*)).mp.

17	(whole genome adj2 (vaccin* or immun* or challenge*)).mp.

18	(experimental adj3 (vaccin* or immuni* or challenge*)).mp.

19	((preclinical or pre-clinical) adj2 study).mp.

20	clinical study/ or controlled clinical trial/ or multicenter study/ or phase 1 clinical trial/ or phase 2 clinical trial/ or phase 3 clinical trial/

21	placebo*.mp.

22	controlled trial.mp.

23	CHIM.mp.

24	controlled human infection.mp.

25	human challenge.mp.

26	Vaccine/

27	Vaccination.mp.

28	(((nonhuman or non-human) adj2 primate*) or NHPs).mp.

29	15 or 16 or 17 or 18 or 19 or 20 or 21 or 22 or 23 or 24 or 25 or 26 or 27 or 28

30	3 and 29

31	6 and 29

32	10 and 29

33	13 and 29

34	14 and 29

35	7 and 29

Cochrane Central Register of Controlled Trials

Issue 2 of 12, February 2024

#253	Ebola virus or ebola fever

#254	MeSH descriptor: [Hemorrhagic Fever, Ebola] explode all trees

#255	Lassa fever or Lassa virus

#256	MeSH descriptor: [Lassa Fever] explode all trees

#257	Nipah virus

#258	MeSH descriptor: [Henipavirus Infections] explode all trees

#259	Rift valley fever

#260	MeSH descriptor: [Rift Valley Fever] explode all trees

#261	Middle East Respiratory Syndrome

#262	MERS

#263	Chikungunya Fever or Chikungunya virus

#264	vaccin* or immuni* or challenge*

#265	placebo or CHIM

#266	nonhuman primate*

#267	#264 or #265 or #266

#268	#253 or #254

#269	#267 and #268

#270	#255 or #256

#271	#270 and #267

#272	#257 or #258

#273	#272 and #267

#274	#259 or #260

#275	#274 and #267

#276	#261 or #262

#277	#276 and #267

#278	#263 and #267

Clinicaltrials.gov

vaccine | Ebola Virus Disease

Also searched for Ebola, Virus, Viral

vaccine | Rift Valley Fever

vaccine | lassa

vaccine | Nipah Virus Infection

vaccine | MERS (Middle East Respiratory Syndrome)

Also searched for Middle East Respiratory Syndrome

vaccine | Chikungunya Fever

Also searched for Chikungunya

Science Citation Index-Expanded, CABI: CAB Abstracts® and Global Health® (Web of Science)

|  | Search Query |
| --- | --- |
| #1 | Ebola virus or Ebola fever (Topic) |
| #2 | Lassa virus or Lassa fever (Topic) |
| #3 | TS=(Nipah virus ) |
| #4 | Nipah virus (Topic) |
| #5 | Rift valley fever (Topic) |
| #6 | Middle East Respiratory Syndrome or MERS (Topic) |
| #7 | Chikungunya (Topic) |
| #8 | ((attenuated or mutant or mutated) near/2 (virus* or pathogen* or strain*)) (Topic) |
| #9 | (whole genome near/2 (vaccin* or immun* or challenge*)) (Topic) |
| #10 | (experimental near/2 (vaccin* or immuni* or challenge*)) (Topic) |
| #11 | ((preclinical or pre-clinical) near/2 study) (Topic) |
| #12 | controlled human infection or CHIM or human challenge (Topic) |
| #13 | ((((nonhuman or non-human) near/2 primate*) or NHPs)) (Topic) |
| #14 | #8 OR #9 OR #10 OR #11 OR #12 OR #13 |
| #15 | #14 AND #1 |
| #16 | #14 AND #2 |
| #17 | #14 AND #3 |
| #18 | #14 AND #5 |
| #19 | #14 AND #6 |
| #20 | #14 AND #7 |

Search terms used on PubMed database on 24th February 2025 following conclusion of review to ensure remained up to date (with no additional studies identified):

(Ebola Hemorrhagic Fever or Ebola virus or Lassa virus or Lassa fever or Nipah virus or Rift Valley Fever or Middle eastern respiratory or MERS* or Chikungunya)

AND

(pseudochallenge* or virus attenuation or attenuated virus or mutant virus or vaccin* or pre-clinical study or controlled trial or CHIM)

| **Search** | **Total number of results** | **Number of duplicates deleted** | **Final number of identified studies** |
| --- | --- | --- | --- |
| **EBOV** | 6242 | 2083 | 4159 |
| **LV** | 911 | 284 | 627 |
| **NiV** | 620 | 138 | 482 |
| **RVFV** | 1197 | 389 | 808 |
| **MERS** | 2517 | 362 | 2155 |
| **CHIKV** | 1591 | 397 | 1194 |

Table S1

Number of studies identified through search strategy. 9425 studies remained after removal of duplication across databases and then 5998 remained after duplication across pathogens and removal of articles identified thorough search terms referencing animal studies.

CHIKV = chikungunya, EBOV = Ebola, LV = Lassa, MERS = Middle East Respiratory Syndrome, NiV = Nipah Virus, RVFV = Rift Valley Fever

### S3 Expanded Table of Data Extracted from all included studies

| Author  & Year | Pathogen | Candidate name | Study Phase | Institution | Purpose of candidate | Mutation from wild-type | Dosage | Number exposed | Rate of recovery of attenuated virus | Method for detection of viraemia | Adverse event incidence | Serious adverse events | Mortality | Placebo cohort size/AE/SAE | Follow-up length | Suitable for CHIM | Availability and Regulatory Requirements |
| --- | --- | --- | --- | --- | --- | --- | --- | --- | --- | --- | --- | --- | --- | --- | --- | --- | --- |
| McClain  1998  (1) | CHIKV | TSI-GSD-218 | Phase 1 | USAMRIID | Experimental Live-attenuated vaccine | Two point-mutations on E2 glycoprotein | 4.4 log_10_ PFU  SC | 74 total  N=36 volunteers already immunised against VEEV  N=38 alphavirus-naïve randomised 1:1 to placebo | 36.8% of alphavirus-naïve cohort | Amplification in cell culture | 0 AEs in the VEEV-vaccinated group  Overall AE rate not reported  Headache 12%  Fever 6%  Myalgia/  Arthralgia 1%  Local AE 6% | Nil | Nil | 19  Overall AE rate not reported but authors report not significantly different to vaccinated cohort | 12 months | Yes | Sold by ATCC  Unlicenced for human use  Investigational new drug status closed |
| Edelman  2000  (2) | CHIKV | TSI-GSD-218 | Phase 2 | University of Maryland  USAMRIID  Salk Institute | Experimental Live-attenuated vaccine | Two point-mutations on E2 glycoprotein | 10^5^ PFU  SC | 59 | Not reported | Not reported | Overall AE rates not reported  58% systemic AE  20% local AE  32% related AE | Nil | Nil | 14  64% AE  29% related AE | 12 months | Yes | Sold by ATCC  Unlicenced for human use  Investigational new drug status closed |
| Hoke  2012  (3) | CHIKV | TSI-GSD-218 | Summary of unpublished Phase 1 studies | United States Army | Experimental Live-attenuated vaccine | Two point-mutations on E2 glycoprotein | 3.1 × 10^5^ PFU/mL  0.5 mL IM  2.75x10^4^ PFU/ml  0.5ml SC | 51 total  (Excluding previously published data)  N=30  N=21 | Not reported | Not reported | Overall rates not reported | Not reported explicitly but assumed nil | Not reported explicitly but assumed nil | 31  Overall AE rate not reported but authors report not significantly different to vaccinated cohort | Not reported | Yes | Sold by ATCC  Unlicenced for human use  Investigational new drug status closed |
| Wressnigg  2020  (4) | CHIKV | VLA1553 | Phase 1 | Valneva | Experimental Live-attenuated vaccine | Deletion of part of *nsP3* gene | 3·2 × 10^3^ TCID_50_ /0·1 mL  3·2 × 10^4^  TCID_50_  /mL  3·2 × 10^5^ TCID_50_ /ml  IM | 120 total  N=31  N=30  N=59 | Not reported explicitly but likely very high given inter-quartile ranges presented | RT-qPCR | 73% total  67.7% low  63.3% med  81.4% high  Related AEs 65%  Related severe AEs 10.8% | 0.8%  1 unrelated polytrauma | Not reported explicitly but assumed nil | Nil | 12 months | Yes | Produced by Valneva  Licenced by FDA and MHRA |
| Schneider  2023  (5) | CHIKV | VLA1553 | Phase 3 | Valneva | Experimental Live-attenuated vaccine | Deletion of part of *nsP3* gene | 1 × 10^4^ TCID_50_ per 0·5 mL  IM | 3093 | Not reported | Not reported | 62.5% | 1.5%  (46/3082)  2 related SAEs (myalgia and SIADH) | Not reported | 1035  44.8% AE  0.8% SAE | 6 months | Yes | Produced by Valneva  Licenced by FDA and MHRA |
| McMahon  2024  (6) | CHIKV | VLA1553 | Phase 3 | Valneva | Experimental Live-attenuated vaccine | Deletion of part of *nsP3* gene | 1 × 10^4^ TCID_50_ per 0·5 mL  IM | 408 | Not reported | Not reported | Any AE 72.5%  Any related AE 60.5%  Local AE 19.4%  Any related severe AE 2.7% | 1.2%  (5/408)  0 related SAEs | Nil | Nil | 6 months | Yes | Produced by Valneva  Licenced by FDA and MHRA |
| Pittman  2016a  (7) | RVFV | MP-12 | Phase 1 | USAMRIID | Experimental Live-attenuated vaccine | Multiple mutations across all three segments of virus | 10^4.4^ PFU SC  10^4.4^ PFU SC  1:10 dilution  1:100 dilution  1:1000 dilution  10^4.7^ PFU SC  10^3.4^ PFU IM (n=6)  10^4.4^ PFU IM | 69 total  N=4  N=22 across different dilutions  N=10  N=6  N=27 | 16.3%  (7 of 43 assessed) | 1 (2.3%) via direct plaque assay  6 (14.0%) by nucleic acid amplification | Overall AE rates not reported  Of 43 vaccines in fully published data:  Headache 25.6%  Malaise 14%  Local AE 9.3% | Nil reported | Nil reported | 13  Overall AE rates not reported but maximum rate of any individual AE = 15.4% | 12 months | Yes | Licence owned by Sabin Vaccine Institute  Unlicenced for human use |
| Pittman  2016b  (8) | RVFV | MP-12 | Phase 2 | USAMRIID | Experimental Live-attenuated vaccine | Multiple mutations across all three segments of virus | 10^5^ PFU IM | 19 | 26.3% | Blind passage of plasma on Vero cells | 89.5%  Headache 58%  Malaise 42.1%  Local AE 68.4% | Nil | Nil | Nil | 12 months  (Some for up to 5 years) | Yes | Licence owned by Sabin Vaccine Institute  Unlicenced for human use |
| Leroux-Roels  2024  (9) | RVFV | hRVFV-4s | Phase 1 | Ghent University  Wageningen Bioveterinary Research | Experimental Live-attenuated vaccine | Split M segment genome | 10^4^ TCID_50_  10^5^ TCID_50_  10^6^ TCID_50_ | 60 total  N=20  N=20  N=20 | 0% | RT-qPCR | Overall AE rate not reported  Headache 47%  Fatigue 47%  Local AE 85% | Nil | Nil | 15  Overall AE rate not reported but maximal rate of individual AE = 13% | 6 months | No | Phase 2 study planned |

Table S2. Studies included after full-text review

AE = Adverse Event. ATCC = American Type Culture Collection. CHIKV = Chikungunya virus. CHIM = Controlled Human Infection Models. IM = Intramuscular. PFU = Plaque forming units. RT-qPCR = quantitative reverse transcription polymerase chain reaction. RVFV = Rift Valley Fever Virus. SAE = Serious Adverse Event. SC = subcutaneously. SIADH= Syndrome of Inappropriate Antidiuretic Hormone secretion. USAMRIID = US Army Medical Research Institute of Infectious Disease
