## Supplementary material for "Attenuated viral strains of priority pathogens for potential use in controlled human infection model studies: A scoping review": PRISMA-ScR checklist

**Preferred Reporting Items for Systematic reviews and Meta-Analyses extension for Scoping Reviews (PRISMA-ScR) Checklist**

| **SECTION** | **ITEM** | **PRISMA-ScR CHECKLIST ITEM** | **REPORTED ON PAGE #** |
| --- | --- | --- | --- |
| **TITLE** | | | |
| Title | 1 | Identify the report as a scoping review. | 1 |
| **ABSTRACT** | | | |
| Structured summary | 2 | Provide a structured summary that includes (as applicable): background, objectives, eligibility criteria, sources of evidence, charting methods, results, and conclusions that relate to the review questions and objectives. | 2 |
| **INTRODUCTION** | | | |
| Rationale | 3 | Describe the rationale for the review in the context of what is already known. Explain why the review questions/objectives lend themselves to a scoping review approach. | 5-6  “Given the lack of precedent with CHIMs in these diseases and the presumed heterogeneity of studies, we have conducted a scoping review to systematically examine the literature for attenuated strains of CEPI priority pathogens that have already been administered to humans.” |
| Objectives | 4 | Provide an explicit statement of the questions and objectives being addressed with reference to their key elements (e.g., population or participants, concepts, and context) or other relevant key elements used to conceptualize the review questions and/or objectives. | 6  “We have conducted a scoping review to systematically examine the literature for attenuated strains of CEPI priority pathogens that have already been administered to humans. This will identify candidates that may be developed into novel CHIMs to facilitate trials for MCMs” |
| **METHODS** | | | |
| Protocol and registration | 5 | Indicate whether a review protocol exists; state if and where it can be accessed (e.g., a Web address); and if available, provide registration information, including the registration number. | 7  “…published in a prospectively registered protocol on the Open Science Framework (<https://osf.io/nu3bf/>).” |
| Eligibility criteria | 6 | Specify characteristics of the sources of evidence used as eligibility criteria (e.g., years considered, language, and publication status), and provide a rationale. | 8  “Inclusion criteria were determined a priori: adult humans (≥ 18 years old); deliberately exposed to a near-whole-genome, attenuated version of any of the following priority viruses: Ebolavirus; Lassa mammarenavirus; Nipah virus; Rift Valley fever phlebovirus; chikungunya virus; or Middle East respiratory syndrome–related coronavirus (CoV).” |
| Information sources* | 7 | Describe all information sources in the search (e.g., databases with dates of coverage and contact with authors to identify additional sources), as well as the date the most recent search was executed. | 7  “A comprehensive literature search was last performed on 24^th^ February 2025 in the Cochrane Central Register of Controlled Trials (CENTRAL, published in the Cochrane Library), MEDLINE (via OVID), Embase (via OVID), Science Citation Index (Web of Science), CAB Abstracts & Global Health (Web of Science) databases. We also searched the WHO International Clinical Trials Registry Platform (ICTRP; apps.who.int/trialsearch/) and ClinicalTrials.gov (https:// clinicaltrials.gov/ct2/home) for trials in progress.” |
| Search | 8 | Present the full electronic search strategy for at least 1 database, including any limits used, such that it could be repeated. | S2 (Supplementary material)  Six search strategies included |
| Selection of sources of evidence† | 9 | State the process for selecting sources of evidence (i.e., screening and eligibility) included in the scoping review. | 8  “Two investigators (DOH and VS) independently screened titles and abstracts using Rayyan (https://www.rayyan.ai/) (37). The first 25 title and abstracts were screened together as a pilot to ensure consistency. No automated tools were used.” |
| Data charting process‡ | 10 | Describe the methods of charting data from the included sources of evidence (e.g., calibrated forms or forms that have been tested by the team before their use, and whether data charting was done independently or in duplicate) and any processes for obtaining and confirming data from investigators. | 9  “The full framework for data extraction is presented in Supplementary Table S2. This was developed iteratively with input from authors expert in CHIM development as the search developed. Data was extracted by a single-author (DOH), recorded using Microsoft Excel, and checked by a second (VS).” |
| Data items | 11 | List and define all variables for which data were sought and any assumptions and simplifications made. | 9 & Table S2  “The primary outcome was confirmation the administered viral strain could subsequently be recovered from participants. The secondary outcome was safety of the mutant viruses. Other outcomes were narratively summarised where reported, namely: author; year; institution; mutation from wild-type; study phase; dosage; sample size; comparator; adverse events (AE)/serious adverse events (SAE); follow-up length; and availability and regulatory requirements.” |
| Critical appraisal of individual sources of evidence§ | 12 | If done, provide a rationale for conducting a critical appraisal of included sources of evidence; describe the methods used and how this information was used in any data synthesis (if appropriate). | 9  “A risk of bias assessment was conducted by a single author (DOH). We used the original Cochrane Collaboration Tool (38) for randomised studies. This tool was the most appropriate because the outcome of interest in our review (viraemia) was not the primary outcome of the studies evaluated and this tool provides a general risk of bias assessment rather than against a particular outcome. Non-randomised studies were assessed using the ROBINS-E tool (39). “ |
| Synthesis of results | 13 | Describe the methods of handling and summarizing the data that were charted. | 10  “Some attenuated strains were investigated in more than one study. In those cases, the methodology and results of those studies are presented together – see Table 1 and Supplementary Table S2. We have provided a descriptive and quantitative (where appropriate) summary for each identified attenuated strain. A background for each pathogen is also presented prior to the description of any identified attenuated strains. “ |
| **RESULTS** | | | |
| Selection of sources of evidence | 14 | Give numbers of sources of evidence screened, assessed for eligibility, and included in the review, with reasons for exclusions at each stage, ideally using a flow diagram. | 10,12  “The literature search resulted in 13,078 studies (n=6242 for EBOV, n=2517 for MERS, n=1591 for CHIKV, n=1197 for RVFV, n=911 for LV and n=620 for NiV). We first removed 3653 duplicates and then, as per our protocol, removed 3427 articles found via search-terms that referenced non-human primates, with the option they could be included later if very limited human data was found (this step was not subsequently required). Thus, 5998 studies remained for title and abstract screening. Of these, 351 manuscripts were selected for full text review and nine were included for data extraction”  PRISMA Flow diagram on Page 12 |
| Characteristics of sources of evidence | 15 | For each source of evidence, present characteristics for which data were charted and provide the citations. | 10, 13  “Of the nine included studies, five were randomised controlled trials (40-44), two were randomised controlled Phase 1 trials with a non-randomised safety or confirmatory cohort (45, 46) and two were non-randomised interventional studies (47, 48). Five of the nine studies investigated for recovery of attenuated virus (40, 44-47).” |
| Critical appraisal within sources of evidence | 16 | If done, present data on critical appraisal of included sources of evidence (see item 12). | 11,14  “Figure 2 presents risk of bias assessments for the included studies. Only one study was found to be at high risk of bias (45). “ |
| Results of individual sources of evidence | 17 | For each included source of evidence, present the relevant data that were charted that relate to the review questions and objectives. | 13 & Table S2 |
| Synthesis of results | 18 | Summarize and/or present the charting results as they relate to the review questions and objectives. | 15-24  Comprehensive results presented for each pathogen |
| **DISCUSSION** | | | |
| Summary of evidence | 19 | Summarize the main results (including an overview of concepts, themes, and types of evidence available), link to the review questions and objectives, and consider the relevance to key groups. | 24-27  “We have identified four such strains across two priority pathogens, although only three strains (TSI-GSD-218 (40, 41, 48) and VLA1553 (42, 43, 46) for CHIKV and MP-12 (45, 47) for RVFV) that produce the required virological response necessary for a CHIM (27). The final identified strain, hRVFV-4s of RVFV, does not cause viraemia in pre-clinical settings (115) nor was viraemia detected in humans despite robust RT-qPCR testing (44). There was insufficient published data to assess the attenuated EBOV strain EBOVΔVP30, although based on the pre-clinical data (126), it is unlikely that it would cause detectable viraemia for use as a primary endpoint within a CHIM. Three of the CEPI priority pathogens, NiV, MERS and LV, have no attenuated strains administered to humans. “ |
| Limitations | 20 | Discuss the limitations of the scoping review process. | 27  “In common with all scoping reviews, our findings are dependent on the quality of the included studies…” |
| Conclusions | 21 | Provide a general interpretation of the results with respect to the review questions and objectives, as well as potential implications and/or next steps. | 27  “In conclusion, there are three attenuated viral strains of two CEPI priority pathogens, CHIKV and RVFV, that have been administered to humans that cause a viraemia and may therefore be amenable to development into a novel CHIM.” |
| **FUNDING** | | | |
| Funding | 22 | Describe sources of funding for the included sources of evidence, as well as sources of funding for the scoping review. Describe the role of the funders of the scoping review. | 28  Funding statement provided |

JBI = Joanna Briggs Institute; PRISMA-ScR = Preferred Reporting Items for Systematic reviews and Meta-Analyses extension for Scoping Reviews.

* Where *sources of evidence* (see second footnote) are compiled from, such as bibliographic databases, social media platforms, and Web sites.

† A more inclusive/heterogeneous term used to account for the different types of evidence or data sources (e.g., quantitative and/or qualitative research, expert opinion, and policy documents) that may be eligible in a scoping review as opposed to only studies. This is not to be confused with *information sources* (see first footnote).

‡ The frameworks by Arksey and O’Malley (6) and Levac and colleagues (7) and the JBI guidance (4, 5) refer to the process of data extraction in a scoping review as data charting*.*

§ The process of systematically examining research evidence to assess its validity, results, and relevance before using it to inform a decision. This term is used for items 12 and 19 instead of "risk of bias" (which is more applicable to systematic reviews of interventions) to include and acknowledge the various sources of evidence that may be used in a scoping review (e.g., quantitative and/or qualitative research, expert opinion, and policy document).

*From:* Tricco AC, Lillie E, Zarin W, O'Brien KK, Colquhoun H, Levac D, et al. PRISMA Extension for Scoping Reviews (PRISMAScR): Checklist and Explanation. Ann Intern Med. 2018;169:467–473. [doi: 10.7326/M18-0850](http://annals.org/aim/fullarticle/2700389/prisma-extension-scoping-reviews-prisma-scr-checklist-explanation).
